# The clinical presentation of avoidant restrictive food intake disorder is largely independent of sex, autism spectrum disorder and anxiety traits

**DOI:** 10.1101/2023.02.12.23285723

**Authors:** Rosie Watts, Tanith Archibald, Pippa Hembry, Maxine Howard, Cate Kelly, Rachel Loomes, Laura Markham, Harry Moss, Alfonce Munuve, Anca Oros, Amy Siddall, Charlotte Rhind, Mohammed Uddin, Zain Ahmad, Rachel Bryant-Waugh, Christopher Hübel

## Abstract

**Importance:** Avoidant restrictive food intake disorder (ARFID) is a newly recognised eating disorder in the Diagnostic and Statistical Manual of Mental Disorders, Fifth Edition and in the International Classification of Diseases, Eleventh Revision which shows great heterogeneity in its clinical presentation.

**Objectives:** Here, we examined the clinical characteristics of ARFID and explored the associations between ARFID symptoms and traits of anxiety. We also investigated whether individuals with ARFID show a different clinical presentation based on their biological sex or comorbid autism spectrum disorder (ASD) diagnosis.

**Design, Setting, and Participants:** We recruited 261 consecutive patients from the specialised ARFID outpatient service at the Maudsley Centre for Child and Adolescent Eating Disorders, Michael Rutter Centre, Maudsley Hospital, London, United Kingdom.

**Main Outcomes and Measures:** The parents of the patients completed the Pica, ARFID, Rumination Disorder – ARFID – Questionnaire (PARDI-AR-Q), the Revised Children’s Anxiety and Depression Scale (RCADS) and reported biological sex of their offspring. Age, height, and weight were obtained from medical records. Clinicians reported on comorbid ASD diagnosis and anxiety traits using the Current View Tool.

**Results:** This cross-sectional study included 261 child and adolescent ARFID patients (133 [51%] female) with a median age of 12.7 years (IQR=9.2 to 15.8). Patients’ BMI-for-age z-scores ranged from −6.75 to 4.07 (median = −1.07, IQR = −2.25 to −0.01).

Patients’ comorbid traits of anxiety had the highest correlations with symptoms on the concern about aversive consequences driver of ARFID: panic disorder correlated with physical feelings of panic and anxiety when eating (*r*=0.53, p=7.74 × 10^−31^) and being afraid to eat (*r*=0.42, p=5.13 × 10^−21^); generalised anxiety correlated with physical feelings of panic and anxiety when eating (*r*=0.44, p=7.72 × 10^−23^); and separation anxiety correlated with avoiding eating situations (*r*=0.36, p=2.01 × 10^−15^). Sensory sensitivity to the appearance of food positively correlated with separation anxiety (*r*=0.40, p=1.52 × 10^−16^) and generalised anxiety (*r*=0.36, p=7.16 × 10^−18^).

The sensory sensitivities (RR = 0.96; 95% CI, 0.85 to 1.09; *P* = .53), lack of interest (RR = 1.14; 95% CI, 1.03 to 1.28; *P* = .02) and concern about aversive consequences (RR = 1.27; 95% CI, 1.03 to 1.56; *P* = .03) drivers were independent of patient sex. Comorbid ASD was reported in 74 (28%) ARFID patients. Their parents reported higher rates of food-related sensory sensitivities (RR = 1.26; 95% CI, 1.09 to 1.45; *P*=0.002) and lack of interest (RR = 1.19; 95% CI, 1.05 to 1.34; *P*=0.006) driving their child’s avoidant and restrictive eating than parents of ARFID patients without ASD (127 [49%]).

**Conclusions and Relevance:** Our study highlights that ARFID patients present with varying combinations and severity of food-related sensory sensitivities, lack of interest and concern about aversive consequences which drive their avoidant and restrictive eating. ARFID does not discriminate between male and female children and adolescents or those with or without ASD. Anxiety and ASD can co-occur with ARFID, and ASD may accentuate food-related sensory sensitivities and lack of interest. Healthcare professionals should be aware of the multi-faceted and heterogenous nature of ARFID; it is important that comprehensive multidisciplinary assessments are administered to sufficiently understand the drivers of the eating behaviour and associated physical health, nutritional, and psycho-social risk and impact.

**Key Points:** *Question:* Are the clinical characteristics of avoidant restrictive food intake disorder (ARFID) associated with biological sex, comorbid autism spectrum disorder (ASD) and anxiety traits in patients who present to a specialist ARFID child and adolescent mental health outpatient service?

*Findings:* In our cross-sectional study, the clinical presentation of ARFID is largely independent of patient sex, comorbid ASD and anxiety.

*Meaning:* Regarding these patient characteristics, we detected no marked differences in our patient sample meaning that all patients should be comprehensively assessed by a multidisciplinary team regardless of comorbidity.

## Introduction

Avoidant restrictive food intake disorder (ARFID) is a feeding and eating disorder recognised since 2013 that presents with substantial heterogeneity across the life span.^1–4^ Individuals with ARFID consume a restricted amount or variety of food which can adversely impact weight, growth, nutrition, and psychosocial functioning, and may impair individual or family well-being. Individuals with ARFID do not restrict or avoid food due to distress about body weight or shape, setting them apart from individuals with anorexia nervosa and bulimia nervosa^5,6^. Three main drivers for ARFID symptomatology are given as examples in the Diagnostic and Statistical Manual of Mental Disorders, Fifth Edition (DSM-5) and International Classification of Diseases 11th Revision (ICD-11): 1) avoidance based on sensory characteristics of food (e.g. taste, texture, appearance); 2) apparent lack of interest in eating and food; and 3) concern about aversive consequences of eating, including, for example, choking or vomiting.^1,3^ A dimensional disorder model of ARFID has been theorised where individuals show heterogeneous presentations with varying symptom severity and combinations.^7–9^

As with all eating disorders, the main treatment recommendation is a form of psycho-behavioural therapy which can usually be delivered on an out-patient basis.^12^ However, at present no empirically tested treatments exist and consequently treatment standards are absent.^13,14^ It is imperative that ARFID patients should be treated by multidisciplinary teams not only to target underlying psychological mechanisms and presenting psychiatric symptoms, but also to address nutritional and physical health complications.^15–19^

The prevalence of ARFID varies depending on the underlying sample.^10^ In a population-based study in Switzerland, 3.2% of school age children had self-reported ARFID with an equal sex distribution: 41% of males and 59% of females.^7^ In a retrospective clinical study across seven adolescent eating disorder clinics in the United States of America and in Canada, 13.8% of patients qualified for an ARFID diagnosis with more female ARFID patients (71%) than males (29%).^11^

Sex differences have been described not only in the occurrence of ARFID, but also within its symptom presentation. In a surveillance study in Canada, male ARFID patients showed more food avoidance caused by sensory sensitivities than females,^20^ which contradicts retrospective clinical findings.^21,22^ Patients with concern about aversive consequences are reported to be more often female than male.^22^

Patients who have food-related sensory sensitivities and experience a lack of interest were found to show nutritional deficiencies or slower growth during childhood. The sensory sensitivities driver commonly co-occurred with a lack of interest in eating or food.^22^ Patients with concern about aversive consequences often had an acute onset with weight loss and comorbid anxiety symptoms.^22^

Individuals with ARFID may present with comorbidities, including gastrointestinal symptoms, anxiety disorders (∼71%), and neurodiversity, such as autism spectrum disorder (ASD; ∼13%).^22,23^ In children and adolescents with ARFID, anxiety disorders are reported to be associated with sensory sensitivities and the concern about aversive consequences.^23^ However, detailed investigations of associations between fear-based anxiety (a core symptom of specific phobias, social anxiety, and panic disorder) and distress-based anxiety (the core symptom of generalised anxiety) are missing. A systematic review of ARFID and ASD showed that the three drivers of ARFID can present in patients with ASD, with food related sensory sensitivities being the most commonly reported.^24^

At present, our knowledge of the heterogeneity in ARFID is limited.^25^ Past research has been constrained by small sample sizes, non-standardised clinical assessment, and retrospective reviews of medical records, categorising patients into one of the three drivers. Here, we report on associations 1) among ARFID symptoms and 2) between different types of anxiety traits and ARFID symptoms. Furthermore, we examine if ARFID symptom presentations differ 3) by sex and 4) comorbid ASD in children and adolescents with ARFID who have been clinically assessed and diagnosed with a standardised procedure at the South London and Maudsley (SLaM) National Health Service (NHS) ARFID outpatient service.

## Methods

### Sample

Our sample comprised 261 child and adolescent ARFID patients (234 full diagnosis, 27 subthreshold) attending the Maudsley Centre for Child and Adolescent Eating Disorders (MCCAED) ARFID outpatient service in London, United Kingdom. This service accepts self-referrals, parent/carer referrals, and professional referrals. Patients were clinically diagnosed with a standardised procedure at first assessment. This study did not have exclusion criteria. We computed each analysis using pairwise complete cases and therefore report the number of participants per analysis.

### Ethics

Our study is a clinical audit and was ethically approved by the appropriate local Child and Adolescent Mental Health Service Clinical Governance Approval committee on the 3^rd^ of June 2021.

### Data collection

We collected the cross-sectional data routinely from parents and clinicians when patients first presented to the outpatient service between November 2019 and July 2022.

### Demographics and anthropometrics

Patient biological sex was reported by parents and their age obtained from medical records. Patient height in metres and weight in kilograms were measured by clinicians at assessment, due to Covid-19 restrictions, 12 (6%) were reported from General Practitioners and 119 (57%) were parent-reported. BMI-for-age z-scores were derived from the UK 1990 (UK90) growth reference data^26–28^ using the R package ‘childsds’.^29^

### ARFID symptomology

Parents completed the parent version of the PARDI-AR-Q.^18,30^ This measure of ARFID symptoms includes questions regarding weight loss, difficulty maintaining weight, slow growth, nutritional deficiencies, reliance on oral supplements or enteral feeding, and psychosocial impairment corresponding to the DSM-5 criteria. Three questions cover each ARFID driver: 1) sensory sensitivities, 2) lack of interest, and 3) concern about aversive consequences. Two questions enquire about the psychosocial impact of the ARFID symptoms on a 0–6 point scale. We calculated three sum scores for each driver and a fourth for psychosocial impairment.

### Anxiety traits

We measured patient anxiety using the Revised Children’s Anxiety and Depression Scale-parent (RCADS-P) questionnaire and the clinician-rated Current View Tool.^31–33^ Parents of children ≥8 years answered the RCADS-P which asks how often the young person experiences thoughts, feelings, or emotions on 0 (never) to 4 (always) scale. We included the RCADS-P subscales separation anxiety, social phobia, generalised anxiety, and panic disorder. On the Current View Tool, clinicians rated symptoms of specific phobia, separation anxiety, social anxiety, generalised anxiety, panic disorder, and agoraphobia on a 4-point scale of none, mild, moderate, and severe. We calculated z-scores for both RCADS-P and Current View (correlations ranged from *r*=.38 to *r*=.63 between both scales). Where available, z-scores from the RCADS-P were included. We included Current View z-scores where there was missing RCADS data (including patients <8 years old) and for specific phobia and agoraphobia as these are not measured on the RCADS.

### Autism spectrum disorder

Clinicians recorded comorbid ASD on the Current View Tool Complexity Factor: pervasive developmental disorder (Asperger’s or ASD). If patients did not have a clinical ASD diagnosis but the clinicians observed ASD features or the patient was awaiting an ASD assessment, clinicians recorded it as ‘suspected’. This resulted in three categories: (no ASD diagnosis (reference category); suspected ASD; and ASD).

### Data analysis

Data were analysed in R, version 4.2.1.^34^ We report means, standard deviations, medians, interquartile ranges, and ranges for continuous variables and frequencies for categorical variables (**Table 1**). We calculated correlations using pairwise complete observations among BMI-for-age z-scores, ARFID symptoms, and anxiety z-scores. We calculated biserial correlations between dichotomous and continuous variables; polychoric correlations between categorical variables; and polyserial correlations between categorical and continuous variables using the ‘hetcor’ function from the ‘polycor’ R package.^35^ We calculated Spearman’s rank correlations between continuous variables using the ‘rcorr’ function from the ‘Hmisc’ R package.^36^

**Table 1.** Demographic and clinical characteristics of child and adolescent avoidant restrictive food intake disorder (ARFID) outpatients.

| **Characteristic** | ***N*** | ***N* = 261<sup>a</sup>** |
| --- | --- | --- |
| **Patient biological sex** | 261 |  |
| Male |  | 128 (49%) |
| Female |  | 133 (51%) |
| **Age in years** | 248 |  |
| Mean (SD) |  | 12.1 (4.1) |
| Median (IQR) |  | 12.7 (9.2 to 15.8) |
| Range |  | 2.9, 17.8 |
| **Comorbid autism spectrum disorder (ASD) diagnosis** | 261 |  |
| No ASD diagnosis |  | 127 (49%) |
| Suspected ASD |  | 60 (23%) |
| ASD |  | 74 (28%) |
| **BMI-for-age z-score** | 242 |  |
| Mean (SD) |  | -1.05 (1.63) |
| Median (IQR) |  | -1.07 (-2.25 to -0.01) |
| Range |  | -6.75, 4.07 |
| **A1: Weight loss or difficulty maintaining weight/growth** | 221 |  |
| No |  | 51 (23%) |
| Yes |  | 170 (77%) |
| **A2: Nutritional deficiency** | 184 |  |
| No |  | 122 (66%) |
| Yes |  | 62 (34%) |
| **A3: Enteral feeding or oral supplement** | 221 |  |
| No |  | 134 (61%) |
| Yes |  | 87 (39%) |
| **A4: Psychosocial impairment** | 191 |  |
| No |  | 52 (27%) |

| Characteristic | *N* | *N* = 261<sup>a</sup> |
| --- | --- | --- |
| Yes |  | 139 (73%) |
| BMI = Body mass index; <sup>a</sup>n(%); SD = standard deviation; IQR = interquartile range |  |  |

To test whether patient sex and comorbid ASD are associated with the ARFID drivers, we regressed the drivers on sex and ASD diagnosis adjusting for age. After inspecting outcome distributions, we tested for overdispersion and zero-inflation and computed negative binomial regressions for the sensory sensitivities and the lack of interest drivers and a zero-inflated negative binomial regression for the concern about aversive consequences driver. To correct for multiple testing, we adjusted our alpha threshold using the Bonferroni method.

## Results

### Demographic and clinical characteristics

Of our ARFID patients, 133 (51%) were female and 128 (49%) male with a median age of 12.7 years (IQR=9.2 to 15.8). Of the young people, 74 (28%) had comorbid ASD and a further 60 (23%) had suspected ASD. The most reported ARFID diagnostic criterion was A1 ‘weight loss or difficulty maintaining weight or growth’ (77%), and the sample had a median BMI-for-age z-score of −1.07 (IQR=−2.25 to −0.01), which means that the BMI value that splits the sample in half was one standard deviation below the population mean. The least common diagnostic criterion reported was nutritional deficiencies (34%). Full descriptive statistics are presented in **Table 1**.

### Correlations

**Figure 1** presents the heterogeneous correlation matrix (eTable 1 in the **Supplement**). The number of pairwise complete observations for each individual correlation ranged from 150 to 241 (eTable 2 in the **Supplement**).

**Figure 1.**
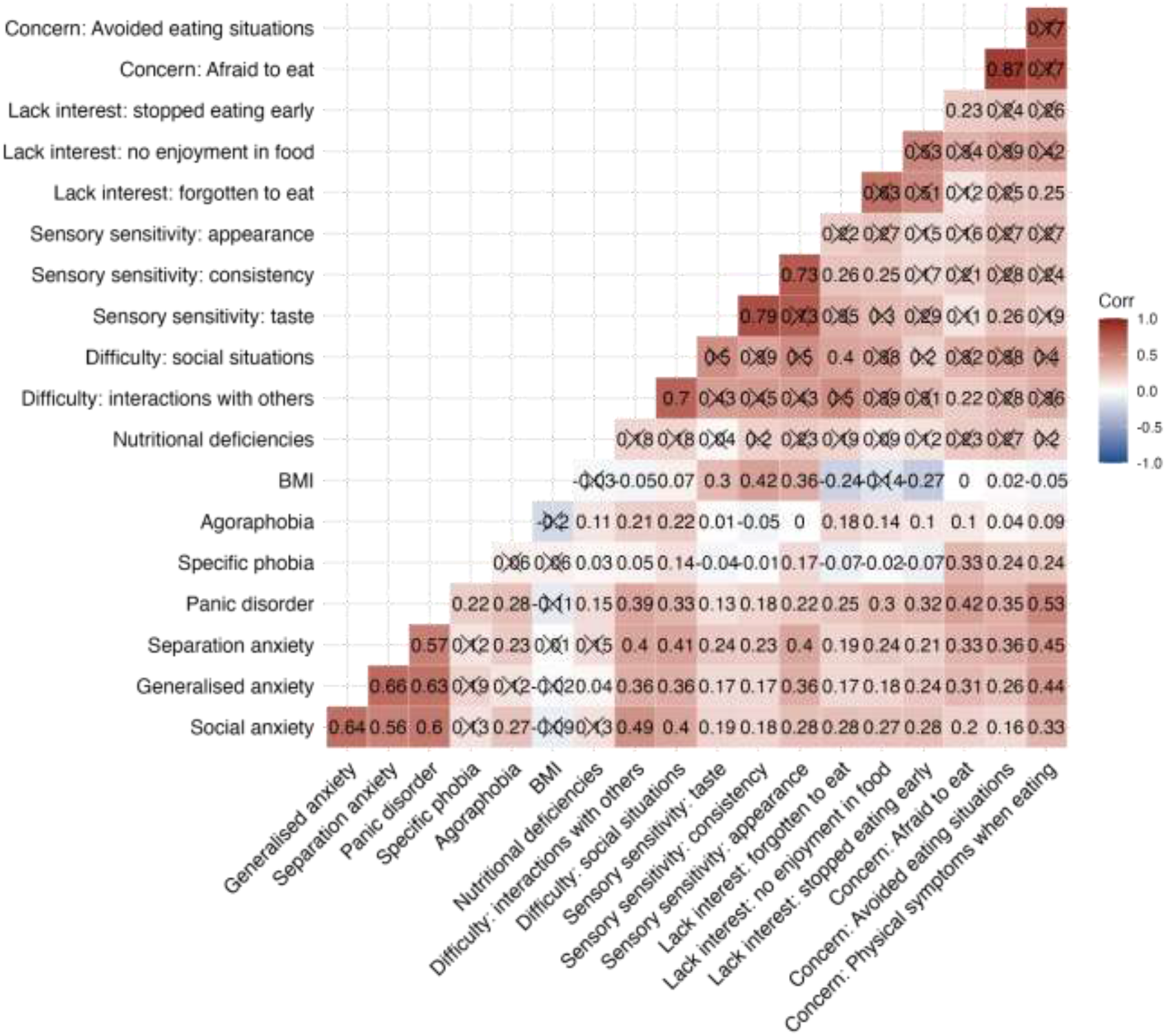
Heterogeneous correlation matrix using pairwise complete observations between symptoms of ARFID measured on the PARDI-AR-Q, BMI-for-age z-scores and comorbid anxiety traits in a clinical sample of children and adolescents ARFID outpatients. BMI = Body mass index; Correlations crossed out did not reach statistical significance at our pre-defined P value threshold of 0.0038 after decomposing the matrix to detect the number of traits and performing Bonferroni adjustment.

Patients’ symptoms on the concern about aversive consequences driver had the highest positive correlation; ‘afraid to eat’ and ‘avoided eating situations’ (*r*=0.87, *P*<2.22 × 10^−16^). The positive correlations between the three sensory sensitivities symptoms of ARFID ranged from 0.73 to 0.79. The lack of interest driver symptoms correlations ranged from 0.51 to 0.63, but they did not reach statistical significance at our predefined *P* value threshold. The positive correlations between symptoms across the three ARFID drivers ranged from 0.11 to 0.42, demonstrating that the drivers are not mutually exclusive.

Patient BMI-for-age z-scores positively correlated with experiencing sensory sensitivities to the taste (*r*=0.30, *P*=1.81 × 10^−14^), consistency (*r*=0.42, *P*=7.41 × 10^−17^), and appearance of food (*r*=0.36, *P*=2.91 × 10^−13^). In contrast, lower BMI-for-age z-scores were associated with patients forgetting to eat (*r*=-0.23, *P*=4.26 × 10^−19^) and stopping eating early (*r*=-0.27, *P*=6.09 × 10^−14^).

Fear and distressed-based anxiety z-scores measured on the RCADS and Current View had the highest correlations with patient symptoms on the concern about aversive consequences driver of ARFID: physical feelings of panic and anxiety when eating with panic disorder (*r*=0.53, *P*=7.74 × 10^−31^), separation anxiety (*r*=0.45, *P*=5.17 × 10^−21^), and generalised anxiety (*r*=0.44, *P*=7.72 × 10^−23^); being afraid to eat with panic disorder (*r*=0.42, *P*=5.13 × 10^−21^); and avoiding eating situations with separation anxiety (*r*=0.36, *P*=2.01 × 10^−15^). Sensory sensitivity to the appearance of food positively correlated with separation anxiety (*r*=0.40, *P*=1.52 × 10^−16^) and generalised anxiety (*r*=0.36, *P*=7.16 × 10^−18^).

### Regressions

We regressed the ARFID drivers on patient sex whilst adjusting for patient age and on ASD diagnosis whilst adjusting for patient age and sex (**Table 2**). Descriptive statistics of the regression analyses are summarised in eTable 3 in the **Supplement**. The sensory sensitivities regression included 217 patients and the lack of interest and concern about aversive consequence regressions included 216 patients each.

**Table 2.** Associations between the clinical drivers of avoidant restrictive food intake disorder and patient’s sex and autism spectrum disorder (ASD), whilst adjusting for age.

| *Explanatory variable* | **Sensory sensitivities** |  |  | **Lack of interest in eating** |  |  | **Concern about aversive consequences of eating** |  |  |
| --- | --- | --- | --- | --- | --- | --- | --- | --- | --- |
|  | *Rate ratio* | *CI (95%)* | *P value<sup>a</sup>* | *Rate ratio* | *CI (95%)* | *P value<sup>a</sup>* | *Rate ratio* | *CI (95%)* | *P value<sup>a</sup>* |
| **Negative Binomial Regressions** |  |  |  |  |  |  |  |  |  |
| ASD diagnosis: suspected<sup>d</sup> | 1.13 | 0.97 to 1.33 | 0.13 | 1.12 | 0.97 to 1.28 | 0.12 | 1.33 | 1.02 to 1.73 | 0.03 |
| ASD diagnosis: ASD<sup>d</sup> | 1.26 | 1.09 to 1.45 | 0.002* | 1.19 | 1.05 to 1.34 | 0.006* | 1.25 | 0.99 to 1.58 | 0.06 |
| Patient sex: female<sup>e</sup> | 0.95 | 0.84 to 1.09 | 0.47 | 1.13 | 1.01 to 1.27 | 0.03 | 1.23 | 0.99 to 1.52 | 0.06 |
| **Zero-Inflated Model** |  |  |  |  |  |  |  |  |  |
| ASD diagnosis: Suspected<sup>d</sup> |  |  |  |  |  |  | 1.11 | 0.48 to 2.57 | 0.81 |
| ASD diagnosis: ASD<sup>d</sup> |  |  |  |  |  |  | 0.73 | 0.32 to 1.67 | 0.45 |
| Patient Sex: Female<sup>e</sup> |  |  |  |  |  |  | 0.62 | 0.30 to 1.26 | 0.19 |
| Observations<sup>b</sup> | 217 |  |  | 216 |  |  | 216 |  |  |
| R<sup>2</sup> Nagelkerke<sup>c</sup> | 0.12 |  |  | 0.152 |  |  | 0.77 / 0.76 |  |  |

CI (95%) = 95% confidence intervals; ASD = autism spectrum disorder; <sup>a</sup>The Bonferroni adjusted significance level is $P=.008$ ; \* indicates statistically significant result <sup>b</sup>Observations are the number of pairwise complete cases per analysis; <sup>c</sup>R<sup>2</sup> Nagelkerke calculated from 'performance' package in R.<sup>37</sup>
<sup>d</sup>The regression models investigating associations between ASD and the ARFID drivers were adjusted for patient sex and age.
<sup>e</sup>The regression models investigating sex differences in the ARFID drivers were adjusted for patient age.

### Sex differences

Male and female patients did not statistically significantly differ in the sensory sensitivities (RR = 0.95; 95% CI, 0.84 to 1.09; *P*= .47), the lack of interest (RR = 1.13; 95% CI, 1.01 to 1.27; *P*= .03) or the concern about aversive consequences (RR = 1.23; 95% CI, .99 to 1.52; *P*= .06) drivers of ARFID. Sex differences in the ARFID drivers are visualised in **Figure 2**.

**Figure 2.**
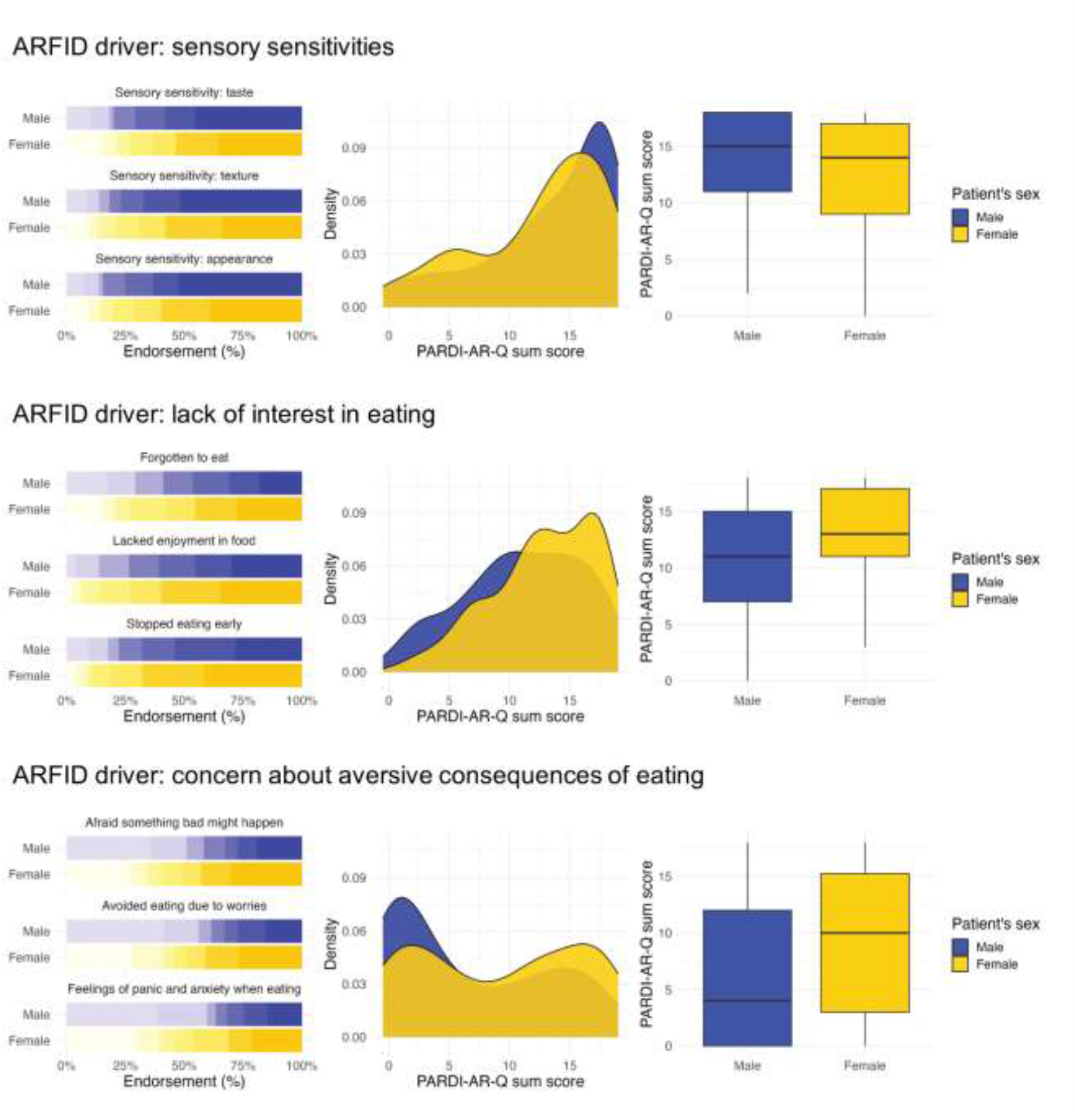
The clinical drivers of avoidant restrictive food intake disorder (ARFID) plotted by sex in child and adolescent ARFID outpatients. **Left**. Bar plots for endorsement of the nine PARDI-AR-Q items measuring the three ARFID drivers (sensory sensitivities, lack of interest, and concern about aversive consequences) on a 0–6-point scale, plotted by patient sex. **Middle**. Density plots for the sum score of the three ARFID drivers plotted by patient sex. **Right**. Box plots for the sum score of the three ARFID drivers plotted by patient sex. PARDI-AR-Q = Pica, ARFID, Rumination Disorder Interview – ARFID – Questionnaire

### ASD

Patients with a comorbid diagnosis of ASD scored on average 26% higher on the sensory sensitivities driver (RR = 1.26; 95% CI, 1.09 to 1.45; *P*=.002) and 19% higher on the lack of interest driver (RR = 1.19; 95% CI, 1.05 to 1.34; *P*=.006) in comparison to the non-ASD group. Comorbid ASD was not associated with the concern about aversive consequences driver (RR = 1.25; 95% CI, 0.99 to 1.58; *P*=.06) or the probability of scoring more than zero on this driver (RR = 0.73; 95% CI, 0.32 to 1.67; *P*=.45). Associations between ASD and the ARFID drivers are visualised in **Figure 3**.

**Figure 3.**
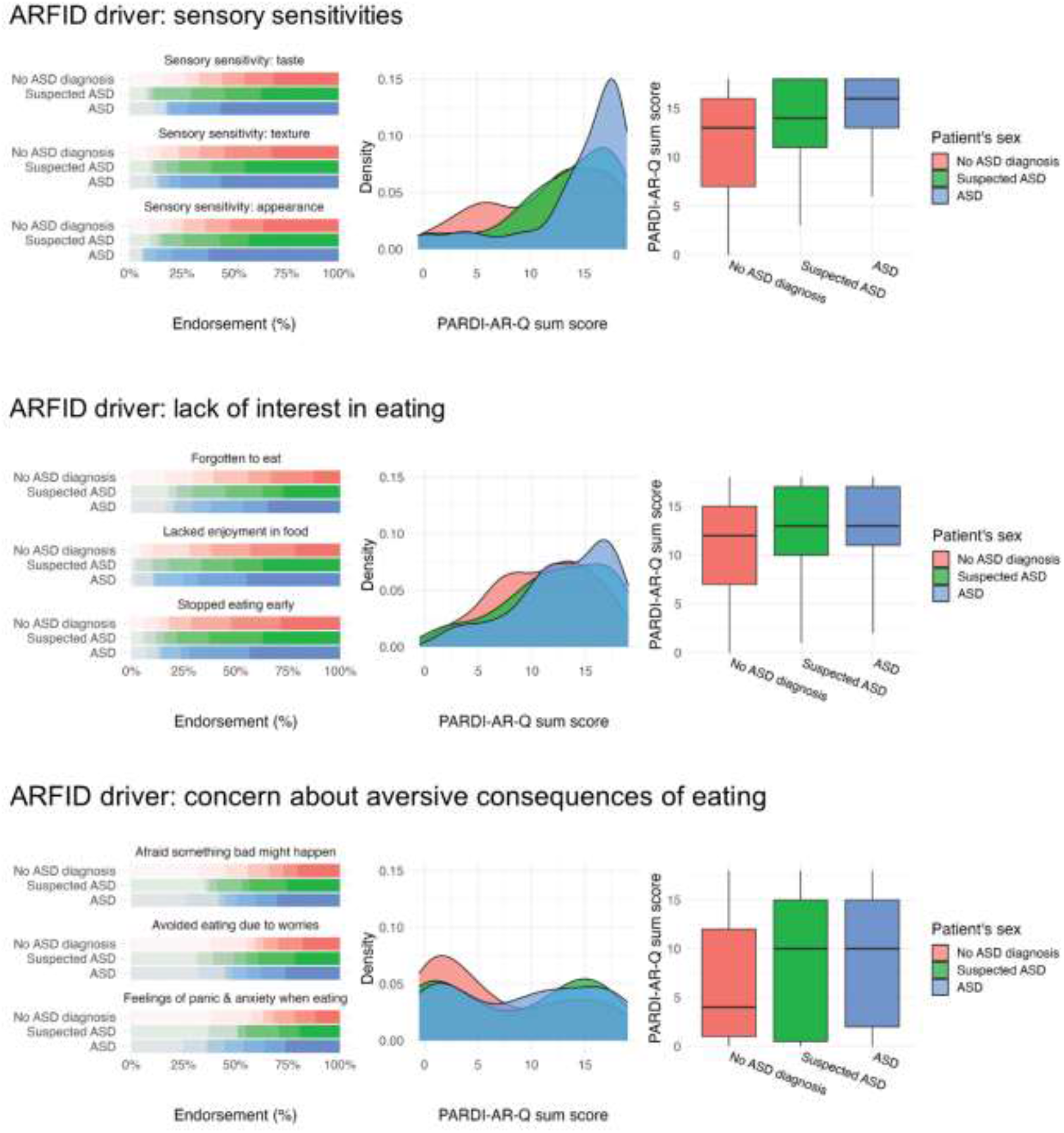
The clinical drivers of avoidant restrictive food intake disorder (ARFID) plotted across comorbid ASD diagnosis in child and adolescent ARFID outpatients. **Left**. Bar plots for endorsement of the nine PARDI-AR-Q items measuring the three ARFID drivers (sensory sensitivities, lack of interest, and concern about aversive consequences) on a 0–6-point scale, plotted by patient ASD diagnosis. **Middle**. Density plots for the sum score of the ARFID drivers, plotted by patient ASD diagnosis. **Right**. Box plots for the sum score of the ARFID drivers, plotted by patient ASD diagnosis. PARDI-AR-Q = Pica, ARFID, Rumination Disorder Interview – ARFID – Questionnaire

## Discussion

ARFID is a relatively newly introduced eating disorder in the DSM-5 and ICD-11 that is hypothesised to be highly heterogenous in its presentation. Our cross-sectional study including 261 ARFID patients aged 2 to 17 years showed that patients experience different severity and combinations of the three main drivers of ARFID: 1) sensory sensitivities, 2) lack of interest in eating and food, and 3) concern about aversive consequences in relation to eating. These drivers contribute to the patient’s avoidant and restrictive eating which in our study was associated with a wide range of BMIs, not only underweight. When we investigated patient characteristics that may be associated with certain symptom constellations, we found that in our sample the ARFID presentation is largely independent of patient sex as none of the drivers showed an association with sex. ARFID can co-occur with psychiatric traits and neurodiversity. In our study, fear- and distress-based anxiety were most strongly associated with the concern about aversive consequences of eating driver and increased sensitivity to the appearance of food. Moreover, our ARFID patients with ASD experienced symptoms of all three ARFID drivers, and they scored significantly higher on the sensory sensitivities and lack of interest drivers than patients without ASD.

During the development of the PARDI-AR-Q, the abovementioned three drivers were identified to play a crucial role in disorder development.^30^ Our correlations between ARFID symptoms measured on the PARDI-AR-Q were congruent with the three drivers of ARFID: sensory sensitivities (*r*=0.73 to *r*=0.79); a lack of interest in eating and food (*r*=0.51 to *r*=0.63); and concern about aversive consequences of eating (*r*=0.77 to *r*=0.87; Figure 1). The symptom correlations further highlighted that the three ARFID drivers do not occur in isolation, providing supporting evidence for a multi-dimensional disorder model of ARFID.^8,38^ Therefore, assuming that the clinical presentation of a patient is only driven by one specific driver is an inappropriate reduction of important clinical information. Clinicians should complete a full multidisciplinary assessment to sufficiently understand the drivers of the eating behaviour and associated physical health, nutritional, and psycho-social risk and impact.

Thomas et al.^39^ theorised a neurobiological model proposing that differences in sensory taste perception, appetite regulation, and attentional bias to fear may be implicated in the aetiology of the three drivers of ARFID. Our patients who had more sensory sensitivities had higher BMI-for-age z-scores, suggesting that these young people may restrict the variety of food consumed rather than the amount.^21,40–42^ Contrastingly, patients who lack interest in eating and food may present with chronic low appetite and early satiety resulting in lower BMIs.^8,43^ This was supported by our findings.

We did not find any specific correlation patterns between fear- or distress-based anxiety traits and ARFID symptoms. Social anxiety, panic disorder and separation anxiety (fear-based anxiety) and generalised (distress-based) anxiety correlated with all symptoms of ARFID. Specific phobia correlated only with symptoms of concern about aversive consequences, which may reflect patients who experience fear of vomiting (emetophobia) or choking.^22,44,45^ The strongest correlations between fear- and distress-based anxiety were with the three ARFID symptoms of concern about aversive consequences and with sensory sensitivities to the appearance of food as assessed by the PARDI-AR-Q. This replicates and extends the finding that anxiety disorders are associated with the concern about aversive consequences and the sensory sensitivities drivers.^23^

Our ARFID patient sample had an equal male-to-female ratio consistent with a population-based study.^7^ Thus, ARFID may be equally common in males and females, differing from anorexia nervosa which affects more females than males.^46–48^ These findings require replication in large epidemiological studies using a two-stage process.

In our sample, patient sex was not significantly associated with any of the three ARFID drivers. Previous research on the symptom presentation reliant on retrospective chart reviews showed that female ARFID patients seem to present with increased concern about aversive consequences compared to males.^22^ In a Canadian Paediatric Surveillance Program, males had more sensory sensitivities than females.^20^ By contrast, in our standardised assessed clinical sample, ARFID presentation did not discriminate between males and females. The difference in findings could be attributed to our clinical sampling process.

In our sample, patients with comorbid ASD showed similar patterns across the three ARFID drivers as patients with suspected ASD or no ASD (Figure 3). Additionally, patients’ comorbid diagnosis of ASD was associated with accentuated sensory sensitivities and lack of interest. Sensory sensitivities, highly-focused special interests and difficulties with interoceptive awareness may be transdiagnostic traits of ASD and ARFID.^49–51^ These traits appear to limit the variety of foods consumed and an individual’s interest in eating and food. By comparison, ARFID patients with and without ASD did not significantly differ on the concern about aversive consequences driver, highlighting that restrictive and repetitive behaviours and interests in patients with ASD do not solely account for eating difficulties.

### Strengths and limitations

Our study benefits from several strengths. We collected one of the largest clinical samples of ARFID patients to date. Data were routinely gathered in a standardised manner when patients first presented to our specialist ARFID outpatient service. Parents reported ARFID symptoms on a dimensional measure capturing the co-occurrence between the three ARFID drivers. Conversely, a few limitations warrant consideration. ARFID symptoms were only reported by parents and future research should collect data from several informants. Due to Covid-19 restrictions during data collection, for 57% of our patients, parents reported anthropometrics. To increase sample size and hence statistical power, we combined data from two measures covering anxiety (i.e., Current View Tool and RCADS) which showed moderate correlations (*r*=.38 to *r*=.63). Furthermore, our study did not consider associations between ARFID presentation and other psychiatric comorbidities, such as depression, obsessive compulsive disorder and attention deficit hyperactivity disorder which should be included in future studies.

## Conclusion

ARFID is a multi-faceted and heterogeneous eating disorder. When assessing young people for ARFID, clinicians should consider that the ARFID drivers are rarely seen in isolation. Our patients presented with a variety of BMIs in the low and high weight range, challenging the common misconception that eating disorders occur solely in individuals who are underweight. Every patient should be comprehensively assessed for all ARFID diagnostic criteria by a multidisciplinary team to establish the physical health, dietetic, and psycho-social risk and impact. Furthermore, multidisciplinary teams can rule out other conditions which may better explain the feeding and eating difficulties. ARFID assessments should involve food diaries reviewed by dietitians to consider nutritional status and to identify patients who may be at high weight yet malnourished. ARFID appears to present similarly across the sexes. Healthcare professionals may increase awareness that males can present with eating disorders, especially ARFID. Anxiety and ASD can co-occur with ARFID, and therefore should be regularly screened for during assessment. ASD can influence ARFID presentation in terms of the strength of some of the drivers of food avoidance and restricted eating, but comorbid ASD is not associated with different drivers. ARFID patients with comorbid ASD should receive the same treatment as those without ASD but may require individualised adaptations to assessment and treatment in light of neurodiversity. Our study raises several important considerations for clinicians assessing and treating young people with avoidant and restrictive eating.

## Data Availability

The code will be available on the following GitHub repository: https://github.com/topherhuebel/arfid

## Acknowledgements

We would like to say thank you to all the children and young people and their families who took part in this research.

## Data availability

The data was routinely gathered during patient treatment and therefore cannot be shared with researchers outside of the National Health Service Trust.

