## Supplementary Material for "The clinical presentation of avoidant restrictive food intake disorder is largely independent of sex, autism spectrum disorder and anxiety traits"

**eTable 1.** Heterogeneous correlation matrix table. Pairwise complete observations between symptoms of Avoidant Restrictive Food Intake Disorder (ARFID) measured on the PARDI-AR-Q, BMI-for-age z-scores and comorbid anxiety traits in a clinical sample of children and adolescents ARFID outpatients.

| Variable 1 | Variable 2 | Correlation coefficient (r) |
|---|---|---|
| Social anxiety | Social anxiety | 1 |
| Generalised anxiety | Social anxiety | 0.640099229 |
| Separation anxiety | Social anxiety | 0.560887547 |
| Panic disorder | Social anxiety | 0.595757023 |
| Specific phobia | Social anxiety | 0.132837961 |
| Agoraphobia | Social anxiety | 0.273324854 |
| BMI | Social anxiety | -0.089733232 |
| Nutritional deficiencies | Social anxiety | 0.131386816 |
| Difficulty: interactions with others | Social anxiety | 0.486836927 |
| Difficulty: social situations | Social anxiety | 0.403329019 |
| Sensory sensitivity: taste | Social anxiety | 0.193595352 |
| Sensory sensitivity: consistency | Social anxiety | 0.183567234 |
| Sensory sensitivity: appearance | Social anxiety | 0.28488235 |
| Lack interest: forgotten to eat | Social anxiety | 0.284410622 |
| Lack interest: no enjoyment in food | Social anxiety | 0.265331488 |
| Lack interest: stopped eating early | Social anxiety | 0.284608359 |
| Fear: Afraid to eat | Social anxiety | 0.195972174 |
| Fear: Avoided eating situations | Social anxiety | 0.157166992 |
| Fear: Physical symptoms when eating | Social anxiety | 0.334380223 |
| Social anxiety | Generalised anxiety | 0.640099229 |
| Generalised anxiety | Generalised anxiety | 1 |
| Separation anxiety | Generalised anxiety | 0.661553505 |
| Panic disorder | Generalised anxiety | 0.629323268 |
| Specific phobia | Generalised anxiety | 0.194275875 |
| Agoraphobia | Generalised anxiety | 0.119866351 |
| BMI | Generalised anxiety | -0.015426102 |
| Nutritional deficiencies | Generalised anxiety | 0.042401717 |
| Difficulty: interactions with others | Generalised anxiety | 0.36395478 |
| Difficulty: social situations | Generalised anxiety | 0.3633476 |
| Sensory sensitivity: taste | Generalised anxiety | 0.169458649 |
| Sensory sensitivity: consistency | Generalised anxiety | 0.173251885 |
| Sensory sensitivity: appearance | Generalised anxiety | 0.359499474 |
| Lack interest: forgotten to eat | Generalised anxiety | 0.172504214 |
| Lack interest: no enjoyment in food | Generalised anxiety | 0.175647449 |
| Lack interest: stopped eating early | Generalised anxiety | 0.237626499 |
| Fear: Afraid to eat | Generalised anxiety | 0.309820692 |
| Fear: Avoided eating situations | Generalised anxiety | 0.264851365 |
| Fear: Physical symptoms when eating | Generalised anxiety | 0.444924185 |
| Social anxiety | Separation anxiety | 0.560887547 |
| Generalised anxiety | Separation anxiety | 0.661553505 |
| Separation anxiety | Separation anxiety | 1 |
| Panic disorder | Separation anxiety | 0.565472138 |

| | | |
|---|---|---|
| Specific phobia | Separation anxiety | 0.123324341 |
| Agoraphobia | Separation anxiety | 0.232402898 |
| BMI | Separation anxiety | 0.010485175 |
| Nutritional deficiencies | Separation anxiety | 0.149730326 |
| Difficulty: interactions with others | Separation anxiety | 0.395771992 |
| Difficulty: social situations | Separation anxiety | 0.410884259 |
| Sensory sensitivity: taste | Separation anxiety | 0.244526581 |
| Sensory sensitivity: consistency | Separation anxiety | 0.22833935 |
| Sensory sensitivity: appearance | Separation anxiety | 0.402889721 |
| Lack interest: forgotten to eat | Separation anxiety | 0.194040627 |
| Lack interest: no enjoyment in food | Separation anxiety | 0.239156331 |
| Lack interest: stopped eating early | Separation anxiety | 0.208297211 |
| Fear: Afraid to eat | Separation anxiety | 0.326888079 |
| Fear: Avoided eating situations | Separation anxiety | 0.356563473 |
| Fear: Physical symptoms when eating | Separation anxiety | 0.453655729 |
| Social anxiety | Panic disorder | 0.595757023 |
| Generalised anxiety | Panic disorder | 0.629323268 |
| Separation anxiety | Panic disorder | 0.565472138 |
| Panic disorder | Panic disorder | 1 |
| Specific phobia | Panic disorder | 0.221014254 |
| Agoraphobia | Panic disorder | 0.278574975 |
| BMI | Panic disorder | -0.10922108 |
| Nutritional deficiencies | Panic disorder | 0.152242934 |
| Difficulty: interactions with others | Panic disorder | 0.389367781 |
| Difficulty: social situations | Panic disorder | 0.331115913 |
| Sensory sensitivity: taste | Panic disorder | 0.131459989 |
| Sensory sensitivity: consistency | Panic disorder | 0.180996057 |
| Sensory sensitivity: appearance | Panic disorder | 0.217648612 |
| Lack interest: forgotten to eat | Panic disorder | 0.25008913 |
| Lack interest: no enjoyment in food | Panic disorder | 0.304677056 |
| Lack interest: stopped eating early | Panic disorder | 0.322963397 |
| Fear: Afraid to eat | Panic disorder | 0.422306141 |
| Fear: Avoided eating situations | Panic disorder | 0.346396086 |
| Fear: Physical symptoms when eating | Panic disorder | 0.531711413 |
| Social anxiety | Specific phobia | 0.132837961 |
| Generalised anxiety | Specific phobia | 0.194275875 |
| Separation anxiety | Specific phobia | 0.123324341 |
| Panic disorder | Specific phobia | 0.221014254 |
| Specific phobia | Specific phobia | 1 |
| Agoraphobia | Specific phobia | 0.064970523 |
| BMI | Specific phobia | 0.055499594 |
| Nutritional deficiencies | Specific phobia | 0.027314865 |
| Difficulty: interactions with others | Specific phobia | 0.045507683 |
| Difficulty: social situations | Specific phobia | 0.142202285 |
| Sensory sensitivity: taste | Specific phobia | -0.03678406 |
| Sensory sensitivity: consistency | Specific phobia | -0.013763385 |
| Sensory sensitivity: appearance | Specific phobia | 0.167951771 |
| Lack interest: forgotten to eat | Specific phobia | -0.068529026 |

| | | |
|---|---|---|
| Lack interest: no enjoyment in food | Specific phobia | -0.022057109 |
| Lack interest: stopped eating early | Specific phobia | -0.071674578 |
| Fear: Afraid to eat | Specific phobia | 0.33346019 |
| Fear: Avoided eating situations | Specific phobia | 0.243007696 |
| Fear: Physical symptoms when eating | Specific phobia | 0.240000008 |
| Social anxiety | Agoraphobia | 0.273324854 |
| Generalised anxiety | Agoraphobia | 0.119866351 |
| Separation anxiety | Agoraphobia | 0.232402898 |
| Panic disorder | Agoraphobia | 0.278574975 |
| Specific phobia | Agoraphobia | 0.064970523 |
| Agoraphobia | Agoraphobia | 1 |
| BMI | Agoraphobia | -0.202005437 |
| Nutritional deficiencies | Agoraphobia | 0.111680498 |
| Difficulty: interactions with others | Agoraphobia | 0.206346847 |
| Difficulty: social situations | Agoraphobia | 0.215496178 |
| Sensory sensitivity: taste | Agoraphobia | 0.008235585 |
| Sensory sensitivity: consistency | Agoraphobia | -0.050240031 |
| Sensory sensitivity: appearance | Agoraphobia | 0.001576084 |
| Lack interest: forgotten to eat | Agoraphobia | 0.177475231 |
| Lack interest: no enjoyment in food | Agoraphobia | 0.144152034 |
| Lack interest: stopped eating early | Agoraphobia | 0.095693684 |
| Fear: Afraid to eat | Agoraphobia | 0.096484555 |
| Fear: Avoided eating situations | Agoraphobia | 0.03852105 |
| Fear: Physical symptoms when eating | Agoraphobia | 0.09423846 |
| Social anxiety | BMI | -0.089733232 |
| Generalised anxiety | BMI | -0.015426102 |
| Separation anxiety | BMI | 0.010485175 |
| Panic disorder | BMI | -0.10922108 |
| Specific phobia | BMI | 0.055499594 |
| Agoraphobia | BMI | -0.202005437 |
| BMI | BMI | 1 |
| Nutritional deficiencies | BMI | -0.026643379 |
| Difficulty: interactions with others | BMI | -0.054381813 |
| Difficulty: social situations | BMI | 0.074511528 |
| Sensory sensitivity: taste | BMI | 0.3010159 |
| Sensory sensitivity: consistency | BMI | 0.41984749 |
| Sensory sensitivity: appearance | BMI | 0.364149812 |
| Lack interest: forgotten to eat | BMI | -0.235949497 |
| Lack interest: no enjoyment in food | BMI | -0.143242095 |
| Lack interest: stopped eating early | BMI | -0.26662547 |
| Fear: Afraid to eat | BMI | 0.003511278 |
| Fear: Avoided eating situations | BMI | 0.023005722 |
| Fear: Physical symptoms when eating | BMI | -0.05156997 |
| Social anxiety | Nutritional deficiencies | 0.131386816 |
| Generalised anxiety | Nutritional deficiencies | 0.042401717 |
| Separation anxiety | Nutritional deficiencies | 0.149730326 |
| Panic disorder | Nutritional deficiencies | 0.152242934 |
| Specific phobia | Nutritional deficiencies | 0.027314865 |

| | | |
|---|---|---|
| Agoraphobia | Nutritional deficiencies | 0.111680498 |
| BMI | Nutritional deficiencies | -0.026643379 |
| Nutritional deficiencies | Nutritional deficiencies | 1 |
| Difficulty: interactions with others | Nutritional deficiencies | 0.180755629 |
| Difficulty: social situations | Nutritional deficiencies | 0.179692402 |
| Sensory sensitivity: taste | Nutritional deficiencies | 0.03651111 |
| Sensory sensitivity: consistency | Nutritional deficiencies | 0.198883167 |
| Sensory sensitivity: appearance | Nutritional deficiencies | 0.229519603 |
| Lack interest: forgotten to eat | Nutritional deficiencies | 0.193878813 |
| Lack interest: no enjoyment in food | Nutritional deficiencies | 0.093236614 |
| Lack interest: stopped eating early | Nutritional deficiencies | 0.124957687 |
| Fear: Afraid to eat | Nutritional deficiencies | 0.229167442 |
| Fear: Avoided eating situations | Nutritional deficiencies | 0.270244879 |
| Fear: Physical symptoms when eating | Nutritional deficiencies | 0.197277409 |
| Social anxiety | Difficulty: interactions with others | 0.486836927 |
| Generalised anxiety | Difficulty: interactions with others | 0.36395478 |
| Separation anxiety | Difficulty: interactions with others | 0.395771992 |
| Panic disorder | Difficulty: interactions with others | 0.389367781 |
| Specific phobia | Difficulty: interactions with others | 0.045507683 |
| Agoraphobia | Difficulty: interactions with others | 0.206346847 |
| BMI | Difficulty: interactions with others | -0.054381813 |
| Nutritional deficiencies | Difficulty: interactions with others | 0.180755629 |
| Difficulty: interactions with others | Difficulty: interactions with others | 1 |
| Difficulty: social situations | Difficulty: interactions with others | 0.696467828 |
| Sensory sensitivity: taste | Difficulty: interactions with others | 0.429751611 |
| Sensory sensitivity: consistency | Difficulty: interactions with others | 0.447478198 |
| Sensory sensitivity: appearance | Difficulty: interactions with others | 0.42712537 |
| Lack interest: forgotten to eat | Difficulty: interactions with others | 0.498378089 |
| Lack interest: no enjoyment in food | Difficulty: interactions with others | 0.388885414 |
| Lack interest: stopped eating early | Difficulty: interactions with others | 0.307270459 |
| Fear: Afraid to eat | Difficulty: interactions with others | 0.221899026 |
| Fear: Avoided eating situations | Difficulty: interactions with others | 0.281521808 |
| Fear: Physical symptoms when eating | Difficulty: interactions with others | 0.356028374 |
| Social anxiety | Difficulty: social situations | 0.403329019 |
| Generalised anxiety | Difficulty: social situations | 0.3633476 |
| Separation anxiety | Difficulty: social situations | 0.410884259 |
| Panic disorder | Difficulty: social situations | 0.331115913 |
| Specific phobia | Difficulty: social situations | 0.142202285 |
| Agoraphobia | Difficulty: social situations | 0.215496178 |
| BMI | Difficulty: social situations | 0.074511528 |
| Nutritional deficiencies | Difficulty: social situations | 0.179692402 |
| Difficulty: interactions with others | Difficulty: social situations | 0.696467828 |
| Difficulty: social situations | Difficulty: social situations | 1 |
| Sensory sensitivity: taste | Difficulty: social situations | 0.503140361 |
| Sensory sensitivity: consistency | Difficulty: social situations | 0.392619407 |
| Sensory sensitivity: appearance | Difficulty: social situations | 0.499125028 |
| Lack interest: forgotten to eat | Difficulty: social situations | 0.397162645 |
| Lack interest: no enjoyment in food | Difficulty: social situations | 0.378420205 |

| | | |
|---|---|---|
| Lack interest: stopped eating early | Difficulty: social situations | 0.197910359 |
| Fear: Afraid to eat | Difficulty: social situations | 0.318711967 |
| Fear: Avoided eating situations | Difficulty: social situations | 0.378358024 |
| Fear: Physical symptoms when eating | Difficulty: social situations | 0.404990833 |
| Social anxiety | Sensory sensitivity: taste | 0.193595352 |
| Generalised anxiety | Sensory sensitivity: taste | 0.169458649 |
| Separation anxiety | Sensory sensitivity: taste | 0.244526581 |
| Panic disorder | Sensory sensitivity: taste | 0.131459989 |
| Specific phobia | Sensory sensitivity: taste | -0.03678406 |
| Agoraphobia | Sensory sensitivity: taste | 0.008235585 |
| BMI | Sensory sensitivity: taste | 0.3010159 |
| Nutritional deficiencies | Sensory sensitivity: taste | 0.03651111 |
| Difficulty: interactions with others | Sensory sensitivity: taste | 0.429751611 |
| Difficulty: social situations | Sensory sensitivity: taste | 0.503140361 |
| Sensory sensitivity: taste | Sensory sensitivity: taste | 1 |
| Sensory sensitivity: consistency | Sensory sensitivity: taste | 0.793956274 |
| Sensory sensitivity: appearance | Sensory sensitivity: taste | 0.72651897 |
| Lack interest: forgotten to eat | Sensory sensitivity: taste | 0.348876087 |
| Lack interest: no enjoyment in food | Sensory sensitivity: taste | 0.298807162 |
| Lack interest: stopped eating early | Sensory sensitivity: taste | 0.285999298 |
| Fear: Afraid to eat | Sensory sensitivity: taste | 0.107840922 |
| Fear: Avoided eating situations | Sensory sensitivity: taste | 0.262652309 |
| Fear: Physical symptoms when eating | Sensory sensitivity: taste | 0.189166466 |
| Social anxiety | Sensory sensitivity: consistency | 0.183567234 |
| Generalised anxiety | Sensory sensitivity: consistency | 0.173251885 |
| Separation anxiety | Sensory sensitivity: consistency | 0.22833935 |
| Panic disorder | Sensory sensitivity: consistency | 0.180996057 |
| Specific phobia | Sensory sensitivity: consistency | -0.013763385 |
| Agoraphobia | Sensory sensitivity: consistency | -0.050240031 |
| BMI | Sensory sensitivity: consistency | 0.41984749 |
| Nutritional deficiencies | Sensory sensitivity: consistency | 0.198883167 |
| Difficulty: interactions with others | Sensory sensitivity: consistency | 0.447478198 |
| Difficulty: social situations | Sensory sensitivity: consistency | 0.392619407 |
| Sensory sensitivity: taste | Sensory sensitivity: consistency | 0.793956274 |
| Sensory sensitivity: consistency | Sensory sensitivity: consistency | 1 |
| Sensory sensitivity: appearance | Sensory sensitivity: consistency | 0.728294571 |
| Lack interest: forgotten to eat | Sensory sensitivity: consistency | 0.262692359 |
| Lack interest: no enjoyment in food | Sensory sensitivity: consistency | 0.253859322 |
| Lack interest: stopped eating early | Sensory sensitivity: consistency | 0.167260062 |
| Fear: Afraid to eat | Sensory sensitivity: consistency | 0.205608389 |
| Fear: Avoided eating situations | Sensory sensitivity: consistency | 0.27844199 |
| Fear: Physical symptoms when eating | Sensory sensitivity: consistency | 0.244963806 |
| Social anxiety | Sensory sensitivity: appearance | 0.28488235 |
| Generalised anxiety | Sensory sensitivity: appearance | 0.359499474 |
| Separation anxiety | Sensory sensitivity: appearance | 0.402889721 |
| Panic disorder | Sensory sensitivity: appearance | 0.217648612 |
| Specific phobia | Sensory sensitivity: appearance | 0.167951771 |
| Agoraphobia | Sensory sensitivity: appearance | 0.001576084 |

| | | |
|---|---|---|
| BMI | Sensory sensitivity: appearance | 0.364149812 |
| Nutritional deficiencies | Sensory sensitivity: appearance | 0.229519603 |
| Difficulty: interactions with others | Sensory sensitivity: appearance | 0.42712537 |
| Difficulty: social situations | Sensory sensitivity: appearance | 0.499125028 |
| Sensory sensitivity: taste | Sensory sensitivity: appearance | 0.72651897 |
| Sensory sensitivity: consistency | Sensory sensitivity: appearance | 0.728294571 |
| Sensory sensitivity: appearance | Sensory sensitivity: appearance | 1 |
| Lack interest: forgotten to eat | Sensory sensitivity: appearance | 0.223158557 |
| Lack interest: no enjoyment in food | Sensory sensitivity: appearance | 0.272936654 |
| Lack interest: stopped eating early | Sensory sensitivity: appearance | 0.154777131 |
| Fear: Afraid to eat | Sensory sensitivity: appearance | 0.16491956 |
| Fear: Avoided eating situations | Sensory sensitivity: appearance | 0.272713658 |
| Fear: Physical symptoms when eating | Sensory sensitivity: appearance | 0.273365806 |
| Social anxiety | Lack interest: forgotten to eat | 0.284410622 |
| Generalised anxiety | Lack interest: forgotten to eat | 0.172504214 |
| Separation anxiety | Lack interest: forgotten to eat | 0.194040627 |
| Panic disorder | Lack interest: forgotten to eat | 0.25008913 |
| Specific phobia | Lack interest: forgotten to eat | -0.068529026 |
| Agoraphobia | Lack interest: forgotten to eat | 0.177475231 |
| BMI | Lack interest: forgotten to eat | -0.235949497 |
| Nutritional deficiencies | Lack interest: forgotten to eat | 0.193878813 |
| Difficulty: interactions with others | Lack interest: forgotten to eat | 0.498378089 |
| Difficulty: social situations | Lack interest: forgotten to eat | 0.397162645 |
| Sensory sensitivity: taste | Lack interest: forgotten to eat | 0.348876087 |
| Sensory sensitivity: consistency | Lack interest: forgotten to eat | 0.262692359 |
| Sensory sensitivity: appearance | Lack interest: forgotten to eat | 0.223158557 |
| Lack interest: forgotten to eat | Lack interest: forgotten to eat | 1 |
| Lack interest: no enjoyment in food | Lack interest: forgotten to eat | 0.629235936 |
| Lack interest: stopped eating early | Lack interest: forgotten to eat | 0.512402249 |
| Fear: Afraid to eat | Lack interest: forgotten to eat | 0.117487871 |
| Fear: Avoided eating situations | Lack interest: forgotten to eat | 0.2503073 |
| Fear: Physical symptoms when eating | Lack interest: forgotten to eat | 0.250719224 |
| Social anxiety | Lack interest: no enjoyment in food | 0.265331488 |
| Generalised anxiety | Lack interest: no enjoyment in food | 0.175647449 |
| Separation anxiety | Lack interest: no enjoyment in food | 0.239156331 |
| Panic disorder | Lack interest: no enjoyment in food | 0.304677056 |
| Specific phobia | Lack interest: no enjoyment in food | -0.022057109 |
| Agoraphobia | Lack interest: no enjoyment in food | 0.144152034 |
| BMI | Lack interest: no enjoyment in food | -0.143242095 |
| Nutritional deficiencies | Lack interest: no enjoyment in food | 0.093236614 |
| Difficulty: interactions with others | Lack interest: no enjoyment in food | 0.388885414 |
| Difficulty: social situations | Lack interest: no enjoyment in food | 0.378420205 |
| Sensory sensitivity: taste | Lack interest: no enjoyment in food | 0.298807162 |
| Sensory sensitivity: consistency | Lack interest: no enjoyment in food | 0.253859322 |
| Sensory sensitivity: appearance | Lack interest: no enjoyment in food | 0.272936654 |
| Lack interest: forgotten to eat | Lack interest: no enjoyment in food | 0.629235936 |
| Lack interest: no enjoyment in food | Lack interest: no enjoyment in food | 1 |
| Lack interest: stopped eating early | Lack interest: no enjoyment in food | 0.527237899 |

| | | |
|---|---|---|
| Fear: Afraid to eat | Lack interest: no enjoyment in food | 0.342291983 |
| Fear: Avoided eating situations | Lack interest: no enjoyment in food | 0.38774448 |
| Fear: Physical symptoms when eating | Lack interest: no enjoyment in food | 0.424461069 |
| Social anxiety | Lack interest: stopped eating early | 0.284608359 |
| Generalised anxiety | Lack interest: stopped eating early | 0.237626499 |
| Separation anxiety | Lack interest: stopped eating early | 0.208297211 |
| Panic disorder | Lack interest: stopped eating early | 0.322963397 |
| Specific phobia | Lack interest: stopped eating early | -0.071674578 |
| Agoraphobia | Lack interest: stopped eating early | 0.095693684 |
| BMI | Lack interest: stopped eating early | -0.26662547 |
| Nutritional deficiencies | Lack interest: stopped eating early | 0.124957687 |
| Difficulty: interactions with others | Lack interest: stopped eating early | 0.307270459 |
| Difficulty: social situations | Lack interest: stopped eating early | 0.197910359 |
| Sensory sensitivity: taste | Lack interest: stopped eating early | 0.285999298 |
| Sensory sensitivity: consistency | Lack interest: stopped eating early | 0.167260062 |
| Sensory sensitivity: appearance | Lack interest: stopped eating early | 0.154777131 |
| Lack interest: forgotten to eat | Lack interest: stopped eating early | 0.512402249 |
| Lack interest: no enjoyment in food | Lack interest: stopped eating early | 0.527237899 |
| Lack interest: stopped eating early | Lack interest: stopped eating early | 1 |
| Fear: Afraid to eat | Lack interest: stopped eating early | 0.234335619 |
| Fear: Avoided eating situations | Lack interest: stopped eating early | 0.241164829 |
| Fear: Physical symptoms when eating | Lack interest: stopped eating early | 0.262488084 |
| Social anxiety | Fear: Afraid to eat | 0.195972174 |
| Generalised anxiety | Fear: Afraid to eat | 0.309820692 |
| Separation anxiety | Fear: Afraid to eat | 0.326888079 |
| Panic disorder | Fear: Afraid to eat | 0.422306141 |
| Specific phobia | Fear: Afraid to eat | 0.33346019 |
| Agoraphobia | Fear: Afraid to eat | 0.096484555 |
| BMI | Fear: Afraid to eat | 0.003511278 |
| Nutritional deficiencies | Fear: Afraid to eat | 0.229167442 |
| Difficulty: interactions with others | Fear: Afraid to eat | 0.221899026 |
| Difficulty: social situations | Fear: Afraid to eat | 0.318711967 |
| Sensory sensitivity: taste | Fear: Afraid to eat | 0.107840922 |
| Sensory sensitivity: consistency | Fear: Afraid to eat | 0.205608389 |
| Sensory sensitivity: appearance | Fear: Afraid to eat | 0.16491956 |
| Lack interest: forgotten to eat | Fear: Afraid to eat | 0.117487871 |
| Lack interest: no enjoyment in food | Fear: Afraid to eat | 0.342291983 |
| Lack interest: stopped eating early | Fear: Afraid to eat | 0.234335619 |
| Fear: Afraid to eat | Fear: Afraid to eat | 1 |
| Fear: Avoided eating situations | Fear: Afraid to eat | 0.873303469 |
| Fear: Physical symptoms when eating | Fear: Afraid to eat | 0.772504249 |
| Social anxiety | Fear: Avoided eating situations | 0.157166992 |
| Generalised anxiety | Fear: Avoided eating situations | 0.264851365 |
| Separation anxiety | Fear: Avoided eating situations | 0.356563473 |
| Panic disorder | Fear: Avoided eating situations | 0.346396086 |
| Specific phobia | Fear: Avoided eating situations | 0.243007696 |
| Agoraphobia | Fear: Avoided eating situations | 0.03852105 |
| BMI | Fear: Avoided eating situations | 0.023005722 |

| | | |
|---|---|---|
| Nutritional deficiencies | Fear: Avoided eating situations | 0.270244879 |
| Difficulty: interactions with others | Fear: Avoided eating situations | 0.281521808 |
| Difficulty: social situations | Fear: Avoided eating situations | 0.378358024 |
| Sensory sensitivity: taste | Fear: Avoided eating situations | 0.262652309 |
| Sensory sensitivity: consistency | Fear: Avoided eating situations | 0.27844199 |
| Sensory sensitivity: appearance | Fear: Avoided eating situations | 0.272713658 |
| Lack interest: forgotten to eat | Fear: Avoided eating situations | 0.2503073 |
| Lack interest: no enjoyment in food | Fear: Avoided eating situations | 0.38774448 |
| Lack interest: stopped eating early | Fear: Avoided eating situations | 0.241164829 |
| Fear: Afraid to eat | Fear: Avoided eating situations | 0.873303469 |
| Fear: Avoided eating situations | Fear: Avoided eating situations | 1 |
| Fear: Physical symptoms when eating | Fear: Avoided eating situations | 0.768779145 |
| Social anxiety | Fear: Physical symptoms when eating | 0.334380223 |
| Generalised anxiety | Fear: Physical symptoms when eating | 0.444924185 |
| Separation anxiety | Fear: Physical symptoms when eating | 0.453655729 |
| Panic disorder | Fear: Physical symptoms when eating | 0.531711413 |
| Specific phobia | Fear: Physical symptoms when eating | 0.240000008 |
| Agoraphobia | Fear: Physical symptoms when eating | 0.09423846 |
| BMI | Fear: Physical symptoms when eating | -0.05156997 |
| Nutritional deficiencies | Fear: Physical symptoms when eating | 0.197277409 |
| Difficulty: interactions with others | Fear: Physical symptoms when eating | 0.356028374 |
| Difficulty: social situations | Fear: Physical symptoms when eating | 0.404990833 |
| Sensory sensitivity: taste | Fear: Physical symptoms when eating | 0.189166466 |
| Sensory sensitivity: consistency | Fear: Physical symptoms when eating | 0.244963806 |
| Sensory sensitivity: appearance | Fear: Physical symptoms when eating | 0.273365806 |
| Lack interest: forgotten to eat | Fear: Physical symptoms when eating | 0.250719224 |
| Lack interest: no enjoyment in food | Fear: Physical symptoms when eating | 0.424461069 |
| Lack interest: stopped eating early | Fear: Physical symptoms when eating | 0.262488084 |
| Fear: Afraid to eat | Fear: Physical symptoms when eating | 0.772504249 |
| Fear: Avoided eating situations | Fear: Physical symptoms when eating | 0.768779145 |
| Fear: Physical symptoms when eating | Fear: Physical symptoms when eating | 1 |

PARDI-AR-Q = Pica, ARFID, Rumination Disorder Interview – ARFID – Questionnaire; BMI = Body Mass Index

**eTable2.** Number of complete pairwise observations between correlations.

| Variables | 1 | 2 | 3 | 4 | 5 | 6 | 7 | 8 | 9 | 10 | 11 | 12 | 13 | 14 | 15 | 16 | 17 | 18 | 19 |
|---|---|---|---|---|---|---|---|---|---|---|---|---|---|---|---|---|---|---|---|
| 1.Social anxiety | 240 | 239 | 239 | 239 | 207 | 207 | 225 | 177 | 212 | 212 | 212 | 212 | 212 | 212 | 212 | 212 | 212 | 212 | 212 |
| 2.Generalised anxiety | 239 | 240 | 239 | 239 | 207 | 207 | 225 | 177 | 212 | 212 | 212 | 212 | 212 | 212 | 212 | 212 | 212 | 212 | 212 |
| 3.Separation anxiety | 239 | 239 | 241 | 240 | 208 | 208 | 226 | 177 | 211 | 212 | 212 | 212 | 212 | 212 | 212 | 211 | 212 | 211 | 211 |
| 4.Panic disorder | 239 | 239 | 240 | 240 | 207 | 207 | 225 | 177 | 211 | 211 | 211 | 211 | 211 | 211 | 211 | 211 | 211 | 211 | 211 |
| 5.Specific phobia | 207 | 207 | 208 | 207 | 209 | 206 | 197 | 151 | 179 | 180 | 180 | 180 | 180 | 180 | 180 | 179 | 180 | 179 | 179 |
| 6.Agoraphobia | 207 | 207 | 208 | 207 | 206 | 209 | 197 | 150 | 180 | 181 | 181 | 181 | 181 | 181 | 181 | 180 | 181 | 180 | 180 |
| 7.BMI | 225 | 225 | 226 | 225 | 197 | 197 | 242 | 176 | 211 | 212 | 212 | 212 | 212 | 212 | 212 | 211 | 212 | 211 | 211 |
| 8.Nutritional deficiencies | 177 | 177 | 177 | 177 | 151 | 150 | 176 | 184 | 184 | 184 | 184 | 184 | 184 | 184 | 184 | 184 | 184 | 184 | 184 |
| 9.Difficulty: interactions with others | 212 | 212 | 211 | 211 | 179 | 180 | 211 | 184 | 221 | 221 | 221 | 221 | 221 | 221 | 221 | 221 | 221 | 221 | 221 |
| 10.Difficulty: social situations | 212 | 212 | 212 | 211 | 180 | 181 | 212 | 184 | 221 | 222 | 222 | 222 | 222 | 222 | 222 | 221 | 222 | 221 | 221 |
| 11.Sensory sensitivity: taste | 212 | 212 | 212 | 211 | 180 | 181 | 212 | 184 | 221 | 222 | 222 | 222 | 222 | 222 | 222 | 221 | 222 | 221 | 221 |
| 12.Sensory sensitivity: consistency | 212 | 212 | 212 | 211 | 180 | 181 | 212 | 184 | 221 | 222 | 222 | 222 | 222 | 222 | 222 | 221 | 222 | 221 | 221 |
| 13.Sensory sensitivity: appearance | 212 | 212 | 212 | 211 | 180 | 181 | 212 | 184 | 221 | 222 | 222 | 222 | 222 | 222 | 222 | 221 | 222 | 221 | 221 |
| 14.Lack interest: forgotten to eat | 212 | 212 | 212 | 211 | 180 | 181 | 212 | 184 | 221 | 222 | 222 | 222 | 222 | 222 | 222 | 221 | 222 | 221 | 221 |
| 15.Lack interest: no enjoyment in food | 212 | 212 | 212 | 211 | 180 | 181 | 212 | 184 | 221 | 222 | 222 | 222 | 222 | 222 | 222 | 221 | 222 | 221 | 221 |
| 16.Lack interest: stopped eating early | 212 | 212 | 211 | 211 | 179 | 180 | 211 | 184 | 221 | 221 | 221 | 221 | 221 | 221 | 221 | 221 | 221 | 221 | 221 |
| 17.Fear: Afraid to eat | 212 | 212 | 212 | 211 | 180 | 181 | 212 | 184 | 221 | 222 | 222 | 222 | 222 | 222 | 222 | 221 | 222 | 221 | 221 |
| 18.Fear: Avoided eating situations | 212 | 212 | 211 | 211 | 179 | 180 | 211 | 184 | 221 | 221 | 221 | 221 | 221 | 221 | 221 | 221 | 221 | 221 | 221 |
| 19.Fear: Physical symptoms when eating | 212 | 212 | 211 | 211 | 179 | 180 | 211 | 184 | 221 | 221 | 221 | 221 | 221 | 221 | 221 | 221 | 221 | 221 | 221 |

BMI = Body Mass Index

**eTable 3.** Descriptive statistics of avoidant restrictive food intake disorder (ARFID) symptoms and age by ASD diagnosis and biological sex of children and adolescent ARFID outpatients.

| | **No ASD diagnosis** | | | **Suspected ASD** | | | **ASD** | | |
|---|---|---|---|---|---|---|---|---|---|
| Characteristic | N | **Male**, N = 57 | **Female**, N = 70 | N | **Male**, N = 32 | **Female**, N = 28 | N | **Male**, N = 39 | **Female**, N = 35 |
| **Avoidance of food based on sensory sensitivities** | 102 | | | 51 | | | 69 | | |
| Mean (SD) | | 12.3 (5.6) | 11.0 (5.3) | | 13.3 (5.5) | 12.9 (4.8) | | 14.9 (4.6) | 14.1 (5.4) |
| Median (IQR) | | 13.0 (8.8 to 18.0) | 12.0 (6.0 to 15.0) | | 15.0 (10.5 to 18.0) | 14.0 (11.0 to 16.2) | | 18.0 (13.0 to 18.0) | 16.0 (14.0 to 18.0) |
| Unknown | | 9 | 16 | | 5 | 4 | | 4 | 1 |
| **Lack of interest in eating and food** | 101 | | | 51 | | | 69 | | |
| Mean (SD) | | 10.0 (4.8) | 11.9 (4.1) | | 11.4 (4.8) | 13.1 (5.0) | | 12.0 (4.9) | 14.1 (3.7) |
| Median (IQR) | | 10.0 (7.0 to 14.0) | 13.0 (9.0 to 15.0) | | 12.0 (9.5 to 15.0) | 14.0 (10.8 to 17.2) | | 13.0 (8.5 to 16.0) | 15.0 (12.0 to 17.0) |
| Unknown | | 10 | 16 | | 5 | 4 | | 4 | 1 |
| **Fear of aversive consequences of eating** | 101 | | | 51 | | | 69 | | |
| Mean (SD) | | 5.1 (5.7) | 8.4 (6.5) | | 7.3 (6.8) | 9.8 (6.9) | | 8.1 (6.6) | 9.8 (6.7) |
| Median (IQR) | | 3.0 (0.0 to 8.0) | 7.5 (3.0 to 14.8) | | 4.0 (0.0 to 14.5) | 10.5 (3.2 to 16.2) | | 9.0 (2.0 to 14.0) | 12.0 (3.2 to 15.8) |
| Unknown | | 10 | 16 | | 5 | 4 | | 4 | 1 |

| | No ASD diagnosis | | | Suspected ASD | | | ASD | | |
|---|---|---|---|---|---|---|---|---|---|
| Characteristic | N | **Male**, N = 57 | **Female**, N = 70 | N | **Male**, N = 32 | **Female**, N = 28 | N | **Male**, N = 39 | **Female**, N = 35 |
| **Patient's age in years** | 122 | | | 56 | | | 70 | | |
| Mean (SD) | | 10.7 (3.9) | 13.5 (3.6) | | 11.2 (4.9) | 12.0 (3.5) | | 12.0 (4.2) | 13.2 (3.9) |
| Median (IQR) | | 10.5 (7.7 to 14.1) | 14.8 (10.4 to 16.2) | | 10.2 (6.6 to 16.2) | 12.7 (9.7 to 14.2) | | 12.8 (8.8 to 15.6) | 13.8 (11.2 to 16.2) |
| Unknown | | 0 | 5 | | 3 | 1 | | 2 | 2 |

ASD = autism spectrum disorder; N = number; SD = standard deviation; IQR = interquartile range
